# Feasibility of Rapid Case Ascertainment for Cancer in East Africa: An Investigation of Community-Representative Kaposi Sarcoma in the Era of Antiretroviral Therapy

**DOI:** 10.1101/2021.05.21.21257178

**Authors:** Aggrey Semeere, Helen Byakwaga, Miriam Laker-Oketta, Esther Freeman, Naftali Busakhala, Megan Wenger, Charles Kasozi, Matthew Ssemakadde, Mwebesa Bwana, Michael Kanyesigye, Philippa Kadama-Makanga, Elyne Rotich, Job Kisuya, Edwin Sang, Kara Wools-Kaloustian, Toby Maurer, Andrew Kambugu, Jeffrey Martin

## Abstract

**BACKGROUND:** Rapid case ascertainment (RCA) refers to the expeditious and detailed examination of patients with a potentially rapidly fatal disease shortly after diagnosis. RCA is frequently performed in resource-rich settings to facilitate cancer research. Despite its utility, RCA is rarely implemented in resource-limited settings and has not been performed for malignancies. One cancer and context that would benefit from RCA in a resource-limited setting is HIV-related Kaposi sarcoma (KS) in sub-Saharan Africa.

**METHODS:** To determine the feasibility of RCA of KS, we searched for all potential newly diagnosed KS among HIV-infected adults attending three community-based facilities in Uganda and Kenya. Searching involved querying of electronic medical records, pathology record review, and notification by clinicians. Upon identification, a team verified eligibility and attempted to locate patients to perform RCA, which included epidemiologic, clinical and laboratory measurements.

**RESULTS:** We identified 593 patients with suspected new KS. Of the 593, 171 were ineligible, mainly because biopsy failed to confirm KS (65%) or KS was not new (30%). Among the 422 remaining, RCA was performed within 1 month for 56% of patients and within 3 months for 65% (95% confidence interval: 59 to 70%). Reasons for not performing RCA included intervening death (47%), inability to contact (44%), refusal/unsuitable to consent (8.3%), and patient re-location (0.7%).

**CONCLUSIONS:** We found that RCA — an important tool for cancer research in resource-rich settings — is feasible for the investigation of community-representative KS in East Africa. Feasibility of RCA for KS suggests feasibility for other cancers in Africa.

---

Rapid case ascertainment (RCA) refers to the thorough research-level evaluation of a patient shortly following diagnosis of a particular condition. While RCA can be performed for any disease, its mainly been used to evaluate potentially rapidly progressive/fatal disease. This is because RCA allows one to perform detailed clinical and laboratory measurements prior to either the demise of the patient or before he/she experiences either a spontaneous or treatment-induced change from his/her condition at time of diagnosis [1, 2]. As such, RCA measurements are critical in facilitating research regarding stage of disease at diagnosis (i.e., a key metric for the effectiveness of early detection efforts), determinants of disease occurrence (i.e., etiology), and determinants of prognosis [3]. Since its origin [1, 4, 5], RCA has predominantly been performed in resource-rich settings, where it is mostly used to study cancer (e.g., pancreatic [6-8]). Despite its potential utility, RCA has rarely been used in resource-limited settings, with the exception of selected infectious diseases [9-13] in which its utility is to decrease spread. To our knowledge, RCA has not been used to study cancer in resource-limited settings, and there are reasons to believe that it may not be as successful as in resource-rich regions. These include delays in return of pathology results; lack of access to therapy and hence rapid fatality; difficulties in reaching patients due to incomplete phone and residential locator information; and potential hesitancy among gravely ill patients to consent for research.

One cancer and resource-limited context in which findings from RCA would be useful to inform policy and facilitate research is HIV-related Kaposi sarcoma (KS) in sub-Saharan Africa. In this region, even with increased antiretroviral therapy (ART) availability for treatment of HIV infection, KS is the fifth most common cancer in all adults [14]. In addition to being common, survival following KS diagnosis in Africa is also poor; one-year mortality is between 20 to 40% [15-18]. Advanced stage of KS at time of diagnosis is often hypothesized to explain this high mortality, but population-level description of KS stage at diagnosis is lacking. Increasing availability of ART in Africa [19], however, suggests that the epidemiology of KS may change as it has in resource-rich settings [20-24]. Wider use of ART may change both stage of KS at diagnosis and survival. Yet, if KS continues to occur despite ART, this begs questions as to why. A thorough evaluation of a community-representative sample of patients with KS in Africa as soon as they are diagnosed — through RCA — would facilitate research on all these questions.

To evaluate the feasibility of performing RCA for cancer in a resource-limited setting, we endeavored to perform RCA among a community-representative sample of new diagnoses of KS in East Africa. We describe the methods required to establish RCA, the successes, and limitations of the process.

## Methods

### Overall design

We developed systems to identify and attempt to perform RCA on all HIV-infected adults with newly diagnosed KS at three East African medical facilities (Figure 1). First, we established a variety of mechanisms to identify all patients with potential new diagnoses of KS at the facilities (Table 1). Second, among these potential cases, we excluded all those who did not meet our eligibility criteria of new onset KS amongst HIV-infected adults. Third, among those who remained, we sought to locate the patients, obtain informed consent and perform RCA.

**Table 1.**
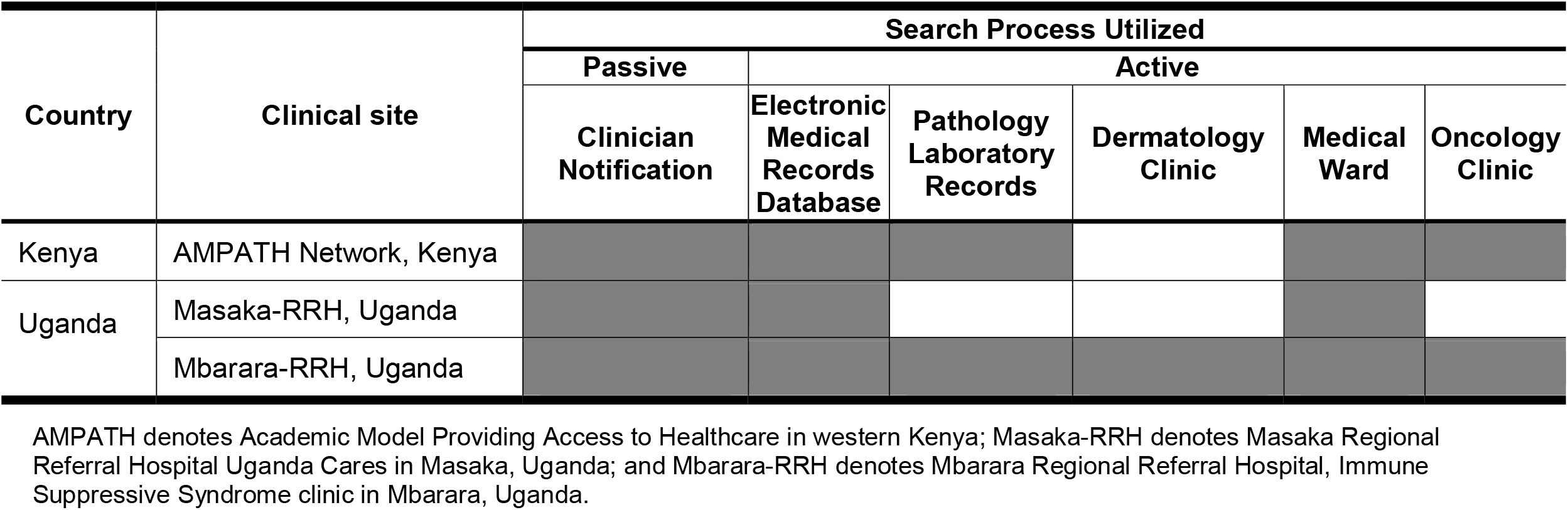
Processes by which patients with possible new Kaposi’s sarcoma were identified at three medical facilities in East Africa. Shaded box denotes use of process at site.

**Figure 1.**
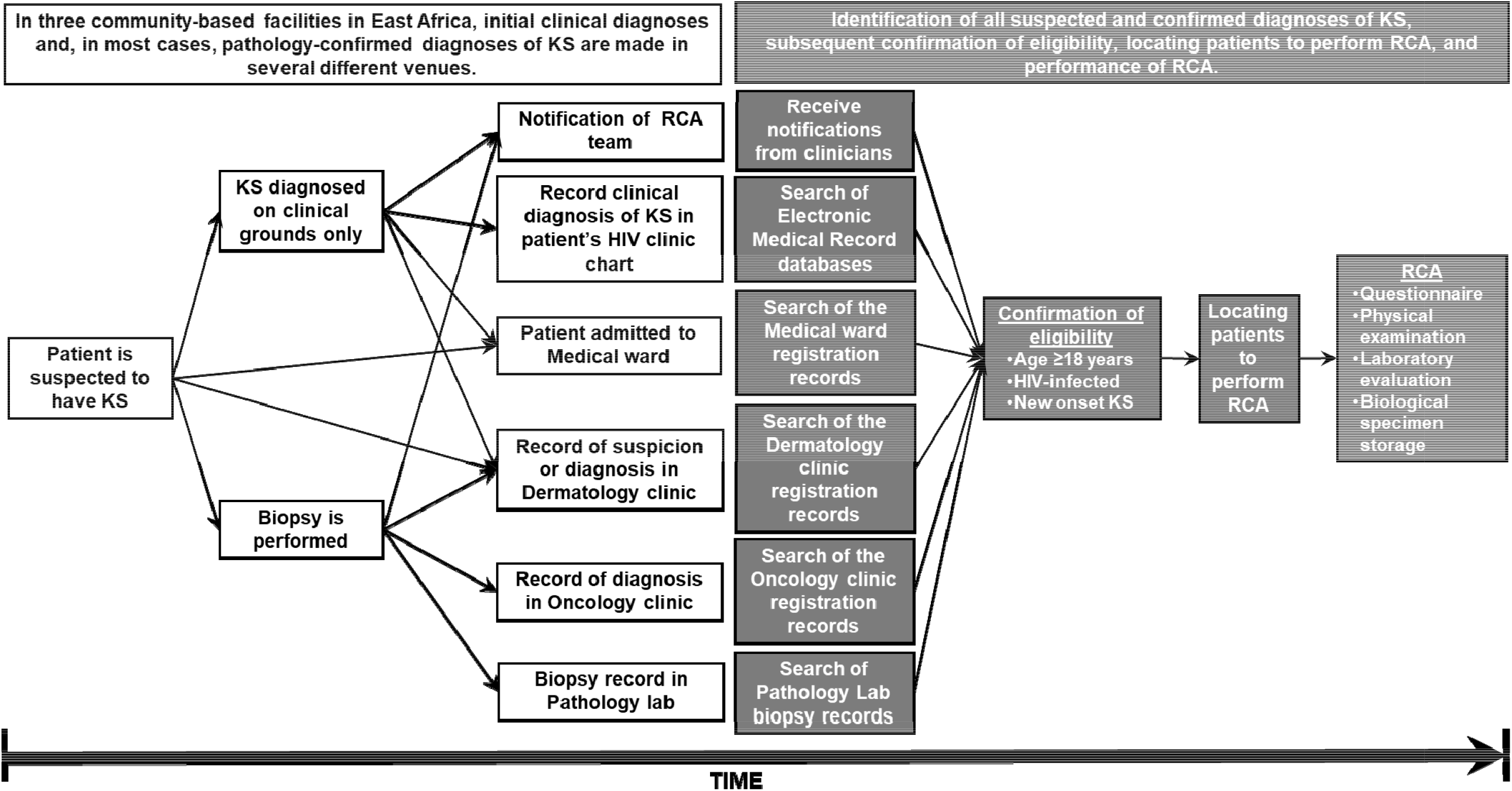
An overview of the process to identify and subsequently perform rapid case ascertainment (RCA) on a community-representative sample of adults with newly diagnosed HIV-related Kaposi sarcoma (KS) at three facilities in East Africa.

### Study sites

We worked at three facilities in East Africa providing primary care to HIV-infected adults: i) The Academic Model Providing Access to Healthcare (AMPATH) network in western Kenya, which has served over 250,000 patients since 2001 and was caring for 70,450 when we began the study in 2016; ii) Masaka Regional Referral Hospital-Uganda Cares clinic (Masaka-RRH), in Masaka, Uganda, which has served over 32,500 patients since 2002 and was caring for 14,380 when we began in 2018; and iii) Mbarara Regional Referral Hospital, Immune Suppressive Syndrome (Mbarara-RRH) clinic in Mbarara, Uganda, which has served over 31,000 patients since 1998 and was caring for 10,800 when we began in 2018.

These sites were chosen because they offered what we believed was the best opportunity to investigate a community-representative sample of new diagnoses of HIV-related KS in East Africa. First, all sites had large primary care populations of HIV-infected adults and inpatient units. Second, all sites had free-of-charge skin biopsies for patients with suspected KS [25]. These services attract referrals not just from existing patients at the facilities but also from those originating outside who may have been out of care or receiving care at facilities without biopsy capability. Third, all sites had electronic medical record (EMR) databases at their HIV primary care clinics that document the patient visit proceedings. Thus, each site, through biopsy availability, offered the best chance in the region to make the diagnosis of KS among community members when KS actually occurred and, through EMR, to centrally assemble all KS diagnoses in preparation for RCA.

### Identification of potential new diagnoses of KS

We used both active and passive mechanisms to search for all potential new diagnoses of KS at the facilities. At each site, we serially queried various fields in the EMR that document any suspicion or confirmation of KS (Table 1). We also manually searched local pathology laboratory records. Patients with a histologic diagnosis of KS or an indeterminate result were, for our purpose of assessing RCA feasibility, considered to have a KS diagnosis. Inpatient adult medical wards and, depending on the site, dermatology and oncology clinics had their registration manually examined for any KS diagnoses (Table 1). For all these active processes, the goal was to perform a weekly or semi-monthly search to identify all KS diagnoses as soon as they occurred. The passive mechanism consisted of obtaining notification (phone or in-person) from clinicians at the facilities regarding any new suspected or biopsy-confirmed case.

### Confirming eligibility

For all patients identified, we evaluated ambient records, awaited pending biopsy results and, where necessary, attempted to call patients or their clinicians in order to confirm our eligibility criteria for pursuit of RCA: HIV-infected; age ≥ 18 years; no prior diagnosis of KS; and either a histologic diagnosis of KS (or, as noted above, an indeterminate result) or a diagnosis on clinical grounds if lesions were deemed unsafe to biopsy (such as ocular lesions).

### Locating patients to perform RCA

We attempted to perform RCA on all those deemed eligible or for whom we could not initially contact to confirm eligibility. An in-person encounter was usually scheduled at the earliest next clinic visit. Occasionally, we approached authorized next-of-kin to arrange the encounter. Failing contact by phone, we would await the patient’s return to care based on scheduling information. If the above failed, we searched for patients in the community using methods we previously established [26].

### Measurements performed during RCA

After providing written informed consent, participants underwent: a) questionnaire-based examination (socio-demographic characteristics, recollection of pathway to KS diagnosis, history of HIV therapy, KS-specific therapy, KS-related symptoms, stigma and quality of life); b) KS-focused physical examination (quantification of anatomic involvement on skin and edema); c) photography of KS lesions; and d) peripheral blood and saliva collection. Peripheral blood was tested for complete blood cell count, CD4+/CD8+ T-cells (FACSCalibur, Becton Dickinson), and plasma HIV RNA viral load (Roche COBAS). Residual blood (stored as plasma, serum, and buffy coat) and saliva were stored at −80°C.

### Statistical Analysis

To describe the feasibility of performing RCA, we performed a time-to-event analysis using the Aalen Johansen estimator [27]. Among patients deemed eligible for us to locate and perform RCA, time zero was the date of health system KS diagnosis (i.e., date of the biopsy result from the pathology lab or date of clinical diagnosis for non-biopsiable KS). The main outcome was performance of RCA. Death without prior RCA was a competing event. All those who did not have RCA performed or not known to be dead were administratively censored at either two years of observation or administrative closure of the analysis, whichever came first. We also evaluated determinants of performing RCA via modified Poisson regression with robust variance [28]. Analyses were performed using Stata (version 16.1, College Station, Texas).

## Results

### Identification of patients with suspected KS

Between July 1, 2016 and April 30, 2019, we identified 593 patients with suspected new KS at three medical facilities in East Africa; 407 (69%) were from AMPATH; 83 (14%) from Masaka-RRH; and 103 (17%) from Mbarara-RRH (Figure 2). Of these, 442 (75%) were identified via clinician notification, 49 (8.3%) via EMR query, 40 (6.7%) via pathology laboratory search, 31 (5.2%) from in-patient wards, 23 (3.9%) from oncology clinic searches, and 8 (1.3%) via dermatology clinic searches. The majority of the clinician notifications were from practitioners on the skin biopsy services.

**Figure 2.**
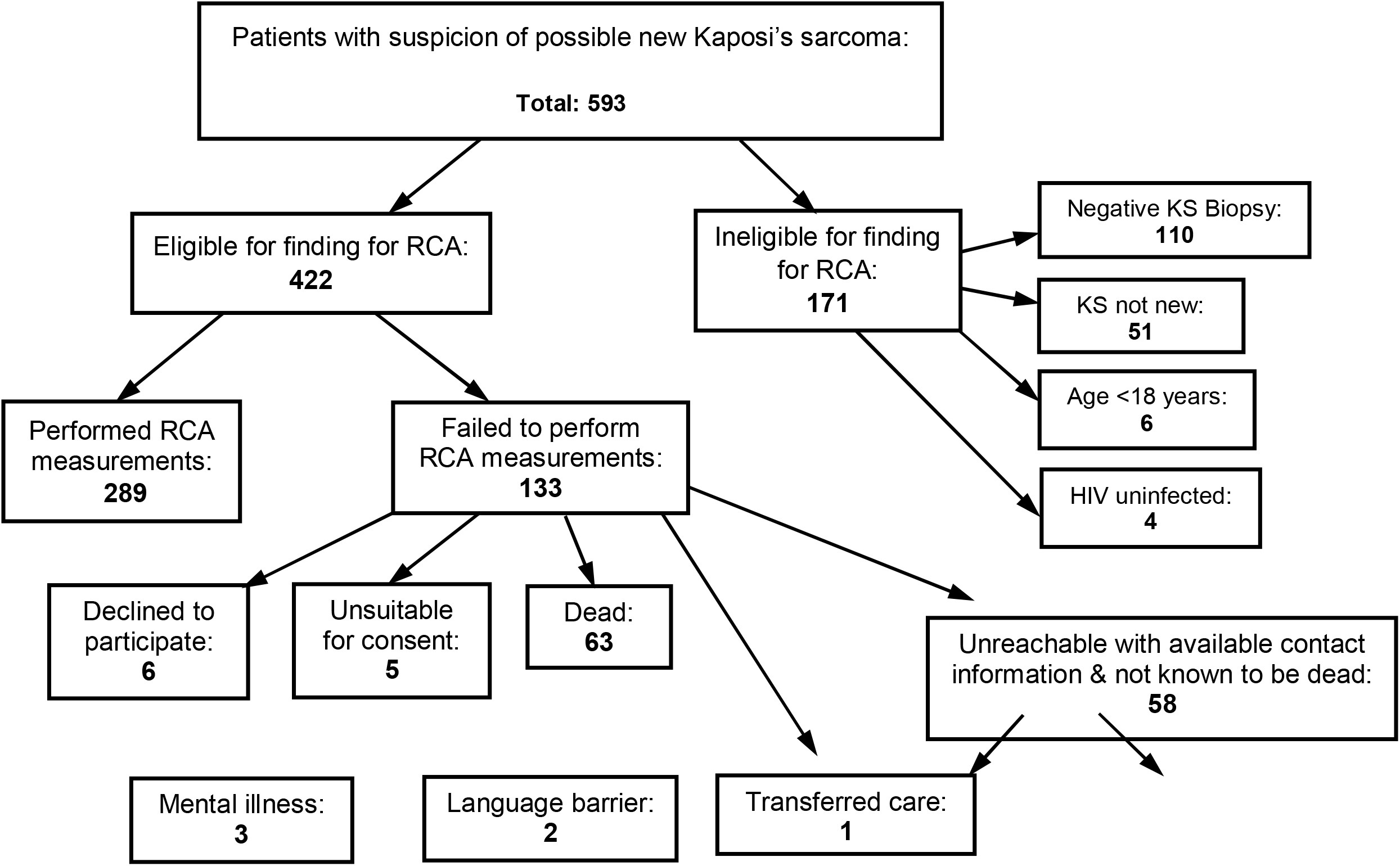
Findings from a process to identify and subsequently perform rapid case ascertainment (RCA) on a community-representative sample of adults with newly diagnosed HIV-related Kaposi sarcoma (KS) at the three facilities in East Africa. AMPATH denotes Academic Model Providing Access to Healthcare in western Kenya; Masaka-RRH denotes Masaka Regional Referral Hospital Uganda Cares in Masaka, Uganda; and Mbarara-RRH denotes Mbarara Regional Referral Hospital, Immune Suppressive Syndrome clinic in Mbarara, Uganda.

### Confirmation of eligibility for RCA

Of the 593 patients identified as potential cases of new KS, 171 were ineligible for further pursuit: 110 (64%) had their skin biopsy rule out KS; 51 (30%) had prior diagnosis of KS; 4 (2.3%) were HIV-uninfected; and 6 (3.5%) were <18 years (Figure 2). The remaining 422 were comprised of 268 (64%) from AMPATH; 77 (18%) from Masaka-RRH, and 77 (18%) from Mbarara-RRH; 202 (34%) were women and median age was 36 years (IQR 31 to 43). Of these 422, 364 (86%) had biopsy-confirmed KS, 7 (1.7%) had an indeterminate pathology result, and 51 (12%) had a lesion clinically compatible with KS but in a location considered unsafe to biopsy.

### Locating patients to perform RCA

Out of 422 patients sought after, we performed RCA among 289. Regarding timing, we performed RCA on 26% (95% confidence interval (CI): 21% to 30%) of the patients within 7 days of health system diagnosis of KS; in 41% within 2 weeks (95% CI: 36% to 46%); in 56% within 1 month (95% CI: 51% to 61%); in 65% within 3 months (95% CI: 59% to 70%); and in 67% within 6 months (95% CI: 61% to 71%) (Figure 3). Intervening death was the main reason for not performing RCA; 60 (11%, 95% CI: 8.0% to 15%) of 422 patients sought after died within 3 months of health system of diagnosis and prior to our ability to perform RCA, and 63 (13%, 95% CI: 9.3% to 17%) died by 2 years. Of those remaining for whom we did not perform RCA and were not known to have died, we located 6 (1.4% of 422) patients who declined to give consent to participate and 1 (0.2%) who had transferred care to a distant region. We also reached 5 patients (1.2%) who were determined unsuitable to provide informed consent because of comprehension or language challenges. The remaining 58 (14%) did not have contact information for tracking in the community or the available information did not enable us to find the patient.

**Figure 3.**
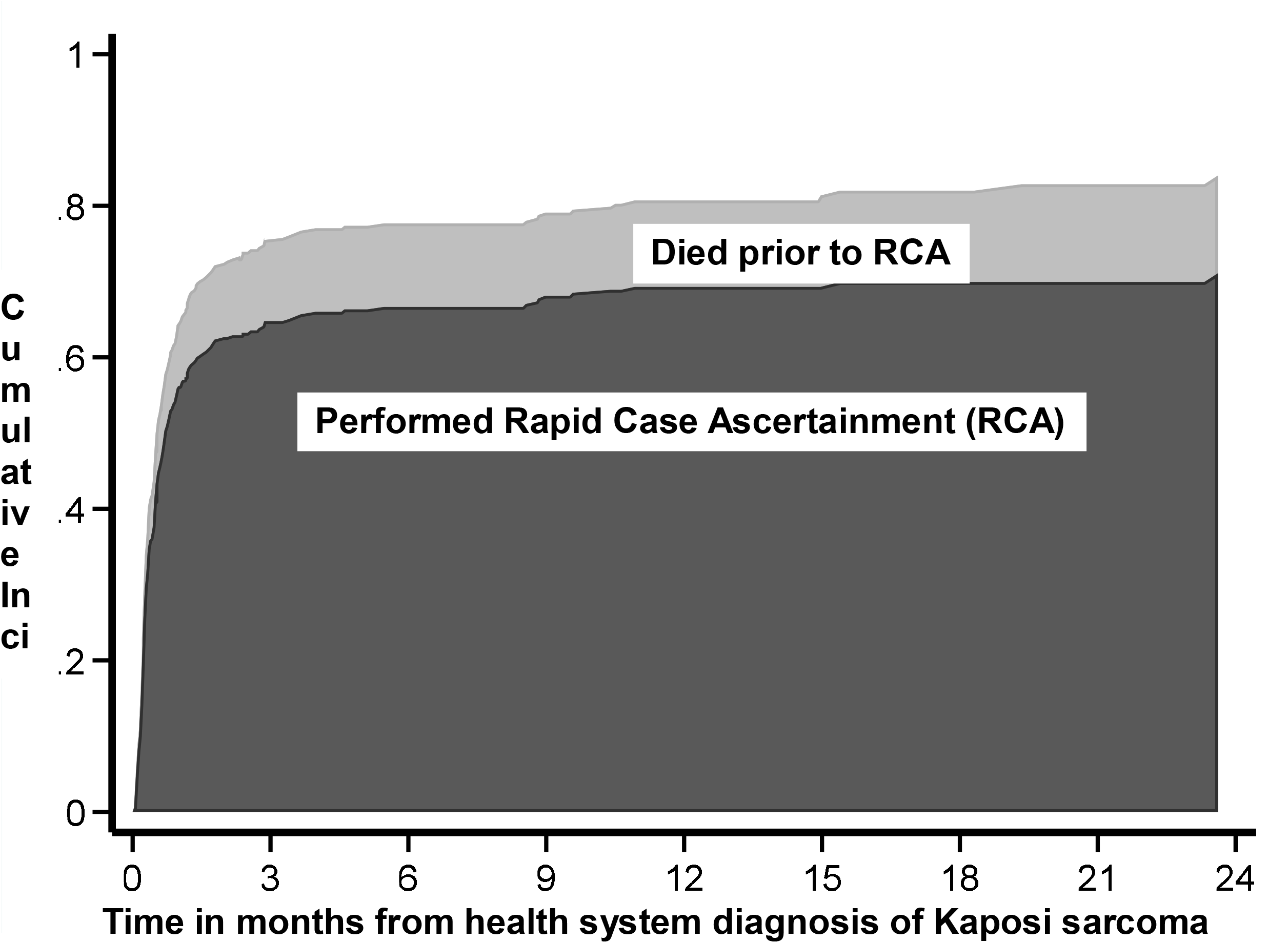
Stacked cumulative incidence plot showing time-to-performing rapid case ascertainment (RCA) (dark grey area) among HIV-infected adults with newly diagnosed Kaposi sarcoma at three medical facilities in East Africa. Cumulative incidence of death prior to RCA (lighter grey area) is shown as a competing event.

Geographic location influenced success in performing RCA. By 3 months following KS diagnosis, RCA was performed amongst 69% (95% CI: 62% to 74%) of patients at AMPATH, 78% (95% CI: 62% to 88%) at Masaka-MRRH; and 45% (95% CI: 33% to 55%) at Mbarara-MRRH. After adjustment for age and sex of the patients, and mode of identification, compared to AMPATH, personnel at Masaka-MRRH were 1.5 times (95% CI: 1.3 to 1.7, *p*<0.001) more likely to perform RCA within 3 months (Table 3). RCA performance also differed by mode of identification. Compared to those identified by clinician notification, patients identified via EMR, or searches of the Oncology clinic, Dermatology clinic, and inpatient medical wards were less likely to have RCA performed at 3 months.

**Table 2.** Demographic, clinical and laboratory characteristics of HIV-infected adult participants newly diagnosed with Kaposi sarcoma at one of three medical facilities in East Africa for whom rapid case ascertainment was performed.

| Characteristic | KS Cases<br>(N=289) |
| --- | --- |
| **Age, years** | 35 (30 to 42)* |
| **Female gender** | 31% |
| **Facility** |  |
| AMPATH, Kenya | 63% |
| Masaka-RRH, Uganda | 26% |
| Mbarara-RRH, Uganda | 12% |
| **Monthly household income, USD** | 120 (50 to 270) |
| **Number of anatomic sites with lesions<sup>†</sup>** | 6 (4 to 10) |
| **Presence of edema<sup>‡</sup>** | 81% |
| **Antiretroviral therapy in use** | 88% |
| **CD4+ T cells, cells/mm<sup>3</sup>** | 246 (97 to 408) |
| **HIV RNA in plasma, copies/ml** |  |
| undetectable (<40) | 43% |
| 40-1,000 | 28% |
| 1,001-10,000 | 8.8% |
| >10,000 | 20% |
| **Hemoglobin, g/dl** | 11 (9 to 13) |
| **ACTG T1 stage<sup>§</sup>** | 91% |
| **ACTG S1 stage<sup>¶</sup>** | 47% |

\* median (interquartile range)
<sup>†</sup> Sites included head, oral cavity, neck, chest, abdomen, back, right upper extremity, left upper extremity, right lower extremity, and left lower extremity. Maximum number of sites is 10.
<sup>‡</sup> Swelling in any of the following sites: faces, genitals, and upper and lower extremities.
<sup>§</sup> One or more of the following: edema; nodular oral lesions; and visceral involvement other than in lymph nodes [1].
<sup>¶</sup> One or more of the following: opportunistic infections; one or more B symptoms; Karnofsky performance status score < 70; and any other HIV-related illness [1]

**Table 3:** Determinants of finding patients diagnosed with Kaposi’s sarcoma to perform rapid case ascertainment within 90 days of health system diagnosis at three clinical sites in sub-Saharan Africa.

|  | Unadjusted |  | Adjusted* |  |
| --- | --- | --- | --- | --- |
|  | Risk Ratio<br>(95% CI) | P value | Risk Ratio<br>(95% CI) | P value |
| **Age of patient, per 1 year increase** | 1.00 (0.99 to 1.004) | 0.44 | 1.00 (0.99 to 1.01) | 0.84 |
| **Sex of patient** |  |  |  |  |
| Women | Ref. |  | Ref. |  |
| Men | 0.98 (0.84 to 1.13) | 0.78 | 0.96 (0.84 to 1.1) | 0.55 |
| **Facility** |  |  |  |  |
| AMPATH, Kenya | Ref. |  | Ref. |  |
| Masaka-RRH, Uganda | 1.4 (1.2 to 1.5) | <0.001 | 1.5 (1.3 to 1.7) | <0.001 |
| Mbarara-RRH, Uganda | 0.66 (0.51 to 0.87) | 0.003 | 0.88 (0.70 to 1.1) | 0.27 |
| **Mode of identification of patient** |  |  |  |  |
| Clinician Notification | Ref. |  | Ref. |  |
| Electronic Medical Records | 0.59 (0.40 to 0.89) | 0.012 | 0.57 (0.39 to 0.85) | 0.006 |
| Pathology Laboratory Records | 1.2 (1.1 to 1.4) | 0.004 | 0.88 (0.73 to 1.1) | 0.18 |
| Dermatology or Oncology Clinic Records | 0.056 (0.0082 to 0.38) | 0.003 | 0.080 (0.012 to 0.54) | 0.010 |
| Medical Ward | 0.61 (0.40 to 0.92) | 0.019 | 0.55 (0.39 to 0.79) | 0.001 |

\* Adjusted risk ratios were derived using modified Poisson regression in a generalized linear model with robust variance and log link function. All variables are adjusted for all variables in the column.
AMPATH denotes Academic Model Providing Access to Healthcare in western Kenya; Masaka-RRH denotes Masaka Regional Referral Hospital Uganda Cares in Masaka, Uganda; and Mbarara-RRH denotes Mbarara Regional Referral Hospital, Immune Suppressive Syndrome clinic in Mbarara, Uganda.

### Description of participants with KS evaluated by RCA

Among 289 participants with KS in whom RCA was performed, 69% were men and median age was 35 years (interquartile range (IQR): 30 to 42) (Table 2). At time of RCA, 88% reported using ART. Regarding the extent of KS disease, participants had a median number of 6 anatomic regions of the skin or mouth affected by KS (IQR: 4 to 10), 81% had leg edema and 91% had AIDS Clinical Trials Group (ACTG) stage T1 KS disease. Laboratory evaluation revealed a median CD4+ T cell count of 246 cells/mm3 (IQR: 97 to 408), and 124 (43%) participants had <40 HIV RNA copies/ml of plasma. Finally, 94% of participants donated peripheral blood and saliva samples for archival storage.

## Discussion

Addressing questions regarding the early detection, etiology and prognosis of particular malignancies requires detailed patient information at time of diagnosis. Much of this information is not collected during routine clinical care and thus requires a dedicated research-level patient encounter. Reaching patients fast enough to record findings that are representative of time of diagnosis is especially challenging with rapidly progressive cancers. RCA was developed for this and is a staple for cancer research in resource-rich settings [1, 4, 7, 8, 29]. In resource-limited settings, the substantive questions and timing challenges are no less relevant, but RCA for cancer has rarely been implemented. We assessed the feasibility — for what we believe is the first time — of RCA for a malignancy in sub-Saharan Africa. We chose to investigate HIV-related KS because its often rapidly progressive course demands RCA, and it has many unanswered substantive questions. We undertook an assessment of RCA feasibility in a context where we had the best chance of studying a community-representative sample of KS, thus yielding more meaningful substantive findings. We were able to locate about two-thirds of adults newly diagnosed with HIV-related KS within 3 months of health system diagnosis, and almost all of these patients consented to provide research-level measurements, including biologic specimens. There were, however, a number of obstacles which limited success and which will require solutions in order to make RCA even more useful.

Previous experience with RCA in sub-Saharan Africa comes from the realm of infectious diseases, specifically, rapidly fatal hemorrhagic fevers [10, 11, 13, 30]. The infectious disease context, however, is considerably different than cancer because diagnostic test results are often returned within one day and more resources are allocated given the public health urgency to isolate the affected patient and protect others [30, 31]. In resource-rich settings, we are not aware of a benchmark optimal pace of RCA for cancer. In one of the earliest reports of large-scale RCA, Wingo et al. endeavored to perform RCA for breast, ovarian and endometrial cancer in a multicenter population-based study in the US [4]. They ascertained 7,010 cancer patients within 8 to 10 weeks, which was a large fraction (about 90%) of their target number of cases. Another account by Beskow et al., summarizing findings from 42 U.S. cancer registries [2], described “rapid” as RCA performed within an average of 44 days with a range of 30 to 90 days after diagnosis. Both reports only describe time to finding patients among those found. This is not the same as the more telling metric that we estimated, which is the cumulative incidence of performing RCA in a given time frame.

While our locating of patients on whom to perform RCA was comparable to resource-rich settings, how we initially identified patients was different. In resource-rich settings, RCA relies on municipally-supported cancer registries to quickly identify all new diagnoses of a given malignancy [2, 6, 32]. Upon identification, registry personnel or interested researchers then track the patient to conduct RCA [4]. While cancer registries exist in East Africa [33], they are currently unsuitable to facilitate RCA either because of unsatisfactory data quality [34] or because of the slow pace of recording diagnoses. Lacking another entity to find the cases, we created several processes to comprehensively scour participating sites for all KS diagnoses. This was facilitated by pre-existing EMRs that could be regularly queried. Importantly, EMR’s include all suspected KS that clinicians deemed unsafe to biopsy and which would be undetected by pathology-based registries. Free-of-charge biopsy services ensured a minimal barrier regarding access to the relevant diagnostic procedure. Collectively, given that a) KS almost always presents with visible skin or mucosal lesions [35]; b) our facilities included several venues in which KS patients receive primary care; c) our sites were well-equipped to diagnose KS when patients presented with suspicious lesions; and d) we were able to learn about most (if not all) diagnoses made at the facilities, we believe that we had a community-representative sample of HIV-related KS available for us to study RCA. The value of a community-representative sample is axiomatic. It allows for scientific inferences generalizable to all KS that occurs in a population, unfettered by selection biases inherent in typical convenience samples in the region that are enriched with patients of sufficient socioeconomic status to be diagnosed or with such extensive disease that it is nearly impossible to escape diagnosis. If short of a community-representative sample, our study population at least represents the burden of KS faced by a community-based health system.

The most common reason for not performing RCA was early death. Among those who died, the median follow-up time was about 6 days, which meant many died even before pathology results became available. Thus, these patients do not so much represent failure of RCA feasibility as they do failure of the diagnostic process. Insufficient contact information was another common explanation for not being able to locate patients. We relied on information routinely recorded at the respective facilities, which was admittedly incomplete and not regularly updated. Because contact information was not collected in anticipation of RCA, its quality may be modifiable in the future. Of note, those patients for whom RCA was not performed because of comprehension or language challenges are also not failures of RCA feasibility but are instead inherent in any research requiring consent.

Performing RCA allowed us to make several initial observations about a community-representative sample of newly diagnosed KS in the era of ART. First, almost all patients had advanced-stage KS. The next step is determining why, specifically whether this is because patients present late for care and/or health systems delay in diagnosing KS. Second, most patients reported to be on ART at the time of RCA, but whether this means that KS developed despite virologically-suppressive ART is unknown. Assuming some patients do develop KS despite virologically-suppressive ART, the next set of questions should address the biologic mechanisms. Third, it is unclear how developing KS despite virologically-suppressive ART will influence survival. For all these questions, having a community-representative sample characterized by RCA is an ideal platform to begin investigation.

There are several limitations that merit mention. Regarding the starting population, despite our extensive efforts to identify all individuals with new KS at our centers, we might have missed some. It is likely those with mild disease were missed, some of which may have resolved on ART alone [36]. In this respect, KS is a challenging cancer to study as it is one of the few where treatment of another condition (in this case HIV) may cure the cancer without the cancer ever being diagnosed. Regarding the representativeness of the group upon which we successfully performed RCA, the large number of deaths that precluded RCA suggests that our RCA group is systematically lacking the most gravely ill patients. Finally, while we observed differences in the success of locating patients to perform RCA by site, we did not collect sufficient process information to understand why. The greater success at one of the sites is proof-of-concept that this level of success is achievable.

There are several implications of our findings. The first is that use of RCA to study HIV-related KS should be expanded in sub-Saharan Africa. With increasing ART use in Africa, KS epidemiology is expected to change. Understanding the public health impact of ART on KS will require documenting stage of KS at diagnosis and survival following diagnosis, both of which are facilitated by RCA. The second implication is that the demonstrated feasibility of RCA for KS in a resource-limited setting suggests it should be promoted for other cancers as well. Yet, to optimize the success of RCA, several conditions are needed. First, stakeholders will require education about RCA. We anecdotally found near-uniform unfamiliarity with RCA; most observers believed it was an intervention intended to improve early detection or treatment. Second, for RCA to yield meaningful inferences, it should be performed on representative populations. Third, prior to beginning an RCA program, systems should be in place to prospectively record and update patient contact information.

In summary, we found that RCA is feasible for the study of HIV-related KS in East Africa, and we demonstrated this using a community-representative sample. Rapid demise of some patients and lack of contact information in others were the main impediments. Success in the study of KS suggests that RCA in other cancers will also be feasible. Cancer epidemiologists in resource-limited settings should begin to include RCA in their repertoire.

## Data Availability

The datasets used and analyzed during the current study are available from the corresponding author on reasonable request.

## Declarations

### Ethics approval and consent to participate

This study had regulatory approval in both Kenya and Uganda. In Kenya the study was approved by the Institutional Research Ethics Committee (IREC) at Moi University in Eldoret, (Protocol No. 0001827) and in Uganda by the Makerere University College of Health Sciences School of Biomedical Sciences Higher Degrees Research and Ethics Committee (Protocol No. SBS-HD-REC-495) and the Uganda National Council of Science and Technology (UNCST) (Protocol No. HS157ES).

### Consent for publication

No individually identifiable data – not applicable.

### Competing Interests

The authors declare that they have no competing interests.

### Funding

Research reported in this publication was supported by the National Institute of Allergy and Infectious Diseases (NIAID) and the National Cancer Institute (NCI) in accordance with the regulatory requirements of the National Institutes of Health under Award Numbers U01AI069911 (East Africa IeDEA Consortium), U54 CA190153, U54 CA254571 and P30 AI027763. The funders did not have a role in design of the study, analysis, interpretation of data, or in writing the manuscript. The content is solely the responsibility of the authors and does not necessarily represent the official views of the National Institutes of Health.

## Acknowledgements

We wish to thank all the research staff: Celestine Lagat, Raphael Kobilo Kipkoir Koima and Linda Chemtai at AMPATH, Eldoret, Kenya; Bronia Mwine, Martin Mwebesa and Placidia Oinembabazi at the ISS Clinic Mbarara Uganda, and Haruna Semuwemba, Stella Nabunya and Fiona Nassonko at Masaka Regional Referral Hospital, Masaka, Uganda who contributed to the collection of data at the different facilities.

## Notes

### Competing Interest Statement

The authors have declared no competing interest.

